# Identification of multi-omic biomarkers from Fecal DNA for improved Detection of Colorectal Cancer and precancerous lesions

**DOI:** 10.1101/2022.11.08.22282099

**Authors:** Yujing Fang, Jiaxi Peng, Zhilong Li, Ruijingfang Jiang, Yuxiang Lin, Ying Shi, Jianlong Sun, Duan Zhuo, Qingjian Ou, Jiali Chen, Xiaohan Wang, Jielun Cai, Shida Zhu, Desen Wan, Yuying Wang, Zhenhai Lu

**Affiliations:** Sun Yat-sen University Cancer Center; State Key Laboratory of Oncology in South China; Collaborative Innovation Center for Cancer Medicine, Guangzhou, China; Envelope Health Biotechnology Co. Ltd, BGI-Shenzhen, Shenzhen, Guangdong, China; BGI Genomics, BGI-Shenzhen, Guangdong, China; Shenzhen Engineering Laboratory for Innovative Molecular Diagnostics, BGI- Shenzhen, Shenzhen, Guangdong, China; BGI-Shenzhen, Shenzhen, Guangdong, China

## Abstract

**Background:** Timely diagnosis and intervention of colorectal cancer (CRC) at curable stages is essential for improving patient survival. Stool samples carry exfoliation of intestinal epithelium, therefore providing excellent opportunity for non-invasive diagnosis of CRC as well as precancerous lesions. In this study, we aimed to conduct multi-dimensional analysis of fecal DNA and investigate the utility of different types of biomarkers for CRC detection.

**Method:** In this case-control study, we performed comprehensive analyses of the genomic, epigenomic, and metagenomic features of fecal DNA from CRC patients, individuals with advanced precancerous lesions (APLs) and controls. DNA methylation markers were identified by whole genome bisulfite sequencing of paired colorectal cancer and normal tissues. A multi-gene fecal DNA methylation test was then developed based on three marker genes (*SDC2, ADHFE1* and *PPP2R5C*) using quantitative methylation-specific PCR (qMSP), and validated on fecal DNA samples. Genomic mutation profiles as well as microbiome signatures of fecal DNA were analyzed using high-throughput sequencing.

**Results:** The methylation-based fecal DNA test demonstrated an overall sensitivity of 88% for CRC and 46.2% for APL respectively, and a specificity of 91.8% for controls. On the other hand, the mutation-based diagnostic model yielded limited sensitivity, and combined detection of methylation markers and mutation in fecal DNA did not improve the assay performance. Meanwhile, a diagnostic model based on the relative abundance of bacterial species showed inferior performance than the methylation-based model. Finally, integrated diagnostic model combining both methylation and microbial markers showed an enhanced performance (AUC= 0.95) compared to methylation markers alone.

**Conclusions:** The multi-gene fecal DNA methylation test provided remarkable diagnostic performance for CRCs and APLs. Furthermore, multi-target assay integrating both methylation and microbial markers may further improve the diagnostic performance. Our findings may aid in the development of novel diagnostic tools for CRC.

## Introduction

Colorectal cancer (CRC) is the third most commonly diagnosed cancer and the second leading cause of cancer death globally^1^. Increased risk of CRC is associated with older age, male gender, lack of physical activity, obesity, unhealthy diet, alcohol consumption, smoking and familial history^2^. The 5-year survival rate approaches nearly 90% in localized CRC but is limited to only 14% in distant metastatic CRC^2, 3^. Unfortunately, CRCs usually present asymptomatic at early stage, leading to a majority of CRC cases (∼60%) being diagnosed when the cancers are metastasized, posing great challenges for treatments and care for the patients^2^.

Most CRCs progress along adenoma-carcinoma sequence, and the entire process is estimated to take as long as 10-15 years^4, 5^. Transition from normal colon epithelium to adenomatous polyps is often accompanied by key somatic mutations such as inactivation of the Adenomatous Polyposis Coli (*APC*) tumor suppressor gene or mutations disrupting β-catenin pathway^6^; this is often followed by acquisition of chromosomal instability and mutations in oncogene *KRAS* and tumor suppressor gene *TP53*, which eventually lead to acquired malignancy^7^. This relatively long-time window provides great opportunity for early detection and timely intervention, i.e., detection and removal of polyps and/or adenoma, which has been shown to effectively reduce CRC incidence^8^. Therefore, screening is key to CRC prevention and it is estimated that more than half of CRC-related death can be prevented through regular screening program^9^. Nevertheless, screening participation rate varies from 16% to 68.2% among different countries and settings due to various reasons including lack of access to screening, limited adherence or/and suboptimal performance when non-invasive test is used for screening^10^. Currently, the most commonly used CRC screening methods are colonoscopy and fecal immunochemical test (FIT) which detects occult blood in stool. Colonoscopy remains the gold standard for CRC screening but it has disadvantages such as being an invasive procedure, associated risk of complications, and being relatively time-consuming. FIT is generally believed to have suboptimal clinical performance (especially for precancerous lesions and early-stage CRC)^11, 12^. The demand for improving current CRC screening strategies necessitates identification of novel biomarkers for CRC and pre-malignant lesions in order to develop cost-effective non-invasive CRC diagnostic and screening tools.

Liquid biopsy using stool, plasma or urine samples is now emerging as a promising tool for cancer detection, screening, diagnosis, and prognosis; being minimally invasive, it allows sampling and profiling of relevant types of cancer, and may overcome tumor heterogeneity compared with tissue biopsy^13^. Of these, stool samples provide excellent opportunity for colorectal cancer detection: exfoliation of normal intestinal epithelium into the colorectal lumen takes place regularly, and this may become more prominent for colorectal neoplasia, which theoretically may be utilized for detection of the neoplasia^14^. Indeed, evidence suggested that genomic and epigenomic alterations accumulated during CRC tumorigenesis could be readily detected from the stool samples of patients^15^. In particular, aberrant methylation of genomic CpG sites appear to arise at early stage during carcinogenesis, making it an attractive biomarker for early detection of malignancy^16, 17^. Previous works have identified a collection of potential CRC-specific methylation markers such as *SFRP2, TFPI2, SDC2, BMP3* and *NDRG4*, etc., and some of these markers have been successfully utilized in commercial diagnostic/screening assays, either used alone, or in combination^18-21^. Additionally, there have also been attempts to integrate multiple types of genomic and/or protein markers into a single detection assay, for example, the Cologuard assay detects *KRAS* mutation, *BMP3* and *NDRG4* methylation and FIT from stool, which achieved a remarkable sensitivity of 92.3% and 42.4% for CRC and advanced adenoma respectively, while having a specificity of 86.6% for non-advanced adenomas or negative findings on colonoscopy^22^. Currently, multiple guidelines endorsed by the American College of Gastroenterology (ACG), the US Preventive Services Task Force (USPSTF) and The American Cancer Society (ACS) recommend this multi-target stool-based assay to be used as an alternative option for CRC screening^23-25^. However, Cologuard remains an expensive assay; also, the relative contribution of the individual biomarker in this multi-target assay remains unclear due to the complexity of the associated algorithm^22^.

Additionally, accumulated evidence has suggested that gut microbiome is closely associated with CRC tumorigenesis^26-31^. Recent evidence also showed that it might be possible to develop stool-based test using CRC or adenoma-enriched microbial markers^32, 33^. It remains to be shown whether these potential microbial markers may be developed into standalone non-invasive test for CRC detection, or used as an assay component to complement performance of fecal DNA-based test.

In the current study, we aimed to comprehensively investigate fecal DNA-derived genomic and epigenomic features that may be utilized for detection of CRC and advanced pre-malignant lesions (APLs). We also identified microbial markers associated with colorectal neoplasia by analyzing the gut microbiome of CRC and adenoma patients as well as control individuals. We were able to develop a methylation-based fecal DNA test that showed high sensitivity and specificity for detection of colorectal neoplasia. We also showed that fecal DNA mutation detection added little to the performance of the methylation-based test; meanwhile, fecal microbiome signature has the potential to be utilized to further improve the performance of methylation-based diagnostic model. Our findings provided valuable insights and may help pave the way for development of more accurate fecal DNA-based CRC diagnostic and/or screening tools in the future.

## Methods

### Study recruitment and sample collection

This study was approved by the Institutional Review Board at Sun Yat-sen University Cancer Center (B2019-068-X01) and BGI (BGI-IRB 20039). Pre-operative CRC patients, high-risk individuals (advanced adenomas, sessile serrate polyps and other high grade epithelial dysplasia) and control individuals with non-advanced neoplasm, negative or non-neoplastic findings on colonoscopy signed written informed consent were enrolled in current study. The inclusion criteria included 1) age between 18 to 80 years old, 2) no personal history of any malignancy, and 3) free of any anti-cancer treatment. Patients diagnosed with other malignancies metastasized to colon or rectum were excluded. Patients with cancers other than colorectal malignancies were recruited as interfering diseases.

38 pairs of colorectal cancer and adjacent normal tissues (NAT) were collected during surgery and immediately frozen at -80°C. 486 stool samples were successfully collected before bowel preparation or five days after colonoscopy without neoplasm resection and stored in a preservation buffer. Homogenized stools were aliquoted and stored at −20°C before DNA extraction.

### Whole genome bisulfite sequencing (WGBS) of tissue samples

A single-stranded DNA (ssDNA) library preparation strategy was adopted to perform WGBS as described previously (cite lung cancer paper). Briefly, gDNA extracted from colorectal cancer and paired NAT tissue was fragmented by sonication. Bisulfite conversion was performed on input DNA using EZ DNA Methylation-Gold™ Kit (Zymo Research, D5006) per manufacturers instructions. Next, bisulfite-converted ssDNA was ligated to sequencing adaptors as described previously. Libraries were sequenced on MGISEQ-2000 using 2×100 paired-end sequencing.

### Identification of differentially methylated lesions (DMLs) and selection of methylation markers

Wald test was performed to identify DMLs from WGBS data of tissue samples based on the Bayesian hierarchical model, with following criteria: methylation ratio difference between tumor and NAT was greater 0.3, and optimal area under the curve (AUC) score was greater than 0.85. DMLs were further selected with the following criteria: AUC > 0.9 for classifying CRC *vs*. NAT tissues; the median methylation ratio in NAT tissues was less than 0.15, and the difference in median methylation ratio between CRC and NAT tissues was more than 0.5. DMLs were annotated using the R package Annotator.

### Quantitative Methylation-Specific Polymerase Chain Reaction (qMSP)

Quantitative methylation-specific polymerase chain reaction (qMSP) assays were designed with Taqman probes for selected DMLs using MethPrimer (www.urogene.org). Assays were first validated using CpGenome Human Methylated & Non-Methylated DNA Standard sets (Merck Millipore, S8001), and assays with satisfactory amplification profile were further validated using tissue and stool samples. For tissue samples, gDNA was isolated using MagPure Buffy Coat DNA Midi KF Kit (Magen, D3537-02). For stool samples, fecal DNA was isolated by Apostle MiniGenomics™ Stool Fast Kit (Apostle, A181206), and 50 μL fecal DNA was used for bisulfite conversion using Sample Pretreatment Kit for Methylation Detection (BGI, EH004).

For the qMSP assay, a total volume of 20 μL of PCR mix composed of 10 ng bisulfite-converted DNA and 200 nM of each primer, 50 nM Taqman probe and 1.25U Hotstart HiTaq DNA Polymerase (Fapon Biotech, MD026). qPCRs were performed on SLAN-96S using a touch-down amplification protocol: 15 s @ 95°C, 10X of (15 s @ 95°C, 30 s @ 65°C to 56°C, 30s @ 72°C), 35X of (15 s @ 95°C, 30 s @ 55°C, 30s @ 72°C).

### Development of Multiplex qMSP Assay

Three methylation markers (*SDC2, ADHFE1* and *PPP2R5C*) that showed the most prominent and distinct methylation signals in CRC tissues *vs*. NAT and passed preliminary validation in a subset of the fecal DNA samples were selected to formulate a multiplexed qMSP assay by applying VIC, FAM, and CY5-labelled probes for the target genes respectively. Primers for internal reference gene (*GAPDH*) were designed to amplify both unmethylated and methylated genomic copies in order to quantify the total human DNA as an internal quality control for successful fecal DNA extraction, bisulfite conversion and amplification. ΔCT value (the Ct value of target gene minus the Ct value of *GAPDH*) was calculated to measure the methylation level of the target genes.

### Mutation Profiling of Fecal DNA

A duplex unique molecular identifier (UMI) strategy was used in library preparation as described previously^34^. Briefly, stool DNA fragmented by sonication was end-repaired and ligated to sequencing adapters, and index PCR was performed, followed by purification with Agencourt AMPure XP beads (Beckman Coulter, A63882).

Target capture reactions were performed using xGen® Lockdown® Reagents (IDT technologies) per manufacturer’s instruction. Captured Libraries were amplified in a 50 μL PCR mix composed of 25 μL 2× KAPA HiFi Hot Start Ready Mix, 5 μL PCR primer pair (10 μM) and 20 μL beads suspensions with the following cycling conditions: 45s at 98°C, followed by 13 cycles of 98°C for 15 s, 60°C for 30 s, and 72°C for 30 s; final extension was performed at 72°C for 1min. Libraries were purified by Agencourt AMPure XP beads, quantified by Qubit™ dsDNA HS Assay Kit, and sequenced on MGISEQ-2000 (MGI Tech) using 2×100 paired-end sequencing.

### Metagenomic Sequencing and Profiling

We performed whole-metagenomic sequencing for 327 fecal samples, using MGISEQ-2000 platform. Paired-end 100-bp reads were quality filtered using FASTP, and then the reads that passed quality filter were aligned to hg38 using bowtie2 to remove human reads. Each sample was down-sampled to 20 million reads randomly and the relative abundance profiles of species were inferred using MetaPhlAn2 with default parameters^35^. In addition, seven public fecal metagenomics datasets of CRCs and controls were obtained from curatedMetagenomicData R package, which used a similar pipeline to preprocess metagenomic data and MetaPhlAn2 to profile species relative abundance.

### Analysis of Microbial Species Abundance and Development of Microbiota-based Diagnostic Model

Linear decomposition model (LDM) was used to analyze association of the species abundance profile with colorectal disease status. Taxa with differential abundance across CRC *vs*. control groups were detected by LDM with false discovery rate (FDR) correction (FDR-adjusted P < 0.01) using the Benjamini-Hochberg procedure, adjusted for age and gender. Results were visualized as a heatmap using the R package ComplexHeatmap.

The relative abundance of all species were used as features and random forest model was fitted using grid search approach with 5-folds cross-validation (Scikit-Learn v0.23.2) to select the best parameters in training set. We used an ensemble of 500 estimator trees; shannon entropy was used to measure the quality of a split at each tree node; and the minimum number of samples per leaf was set to two. To integrate methylation test result and metagenomic data, methylation status of *ADHFE1, PPP2R5C, and SDC2* were incorporated as features (set to 1 if methylation was detected, else 0). Feature importance was obtained from the random forest model training with optimized parameters, and the top-ranking species were utilized in feature reduction modeling.

## Results

### Study design and participants

We performed comprehensive analysis of the genomic, epigenomic, and metagenomic features of colorectal cancer and adenoma patients in this case-control study. Patients diagnosed with colorectal cancer (CRC) (n = 175), advanced precancerous lesion (APL) (n = 26), non-advanced neoplasm (NAN) (n = 65) were included in the study. Individuals with negative or non-neoplastic findings on colonoscopy or other bowel disorders (n = 180), as well as patients with cancers other than colorectal malignancies (n=22) were also included. The detailed information of participants is listed in Table 1.

**Table 1.** Clinical characteristics of the participants included in the study.

|  |  | CRC<br>(n=175) | APL<br>(n=26) | NAN<br>(n=65) | N<br>(n=180) | Interfering<br>Diseases<br>(n=22) |
| --- | --- | --- | --- | --- | --- | --- |
| Age | Median± SD | 56 ± 10 | 65 ± 9 | 69 ± 11 | 63 ± 10 | 57 ± 12 |
| Gender, *n* (%) | Male | 102 (58.3) | 9 (34.6) | 37 (56.9) | 78 (43.3) | 11 (50.0) |
|  | Female | 73 (41.7) | 17 (65.4) | 28 (43.1) | 102 (56.7) | 11 (50.0) |
| Stage, *n* (%) | I | 37 (21.1) |  |  |  |  |
|  | II | 69 (39.4) |  |  |  |  |
|  | III | 54 (30.9) |  |  |  |  |
|  | IV | 15 (8.6) |  |  |  |  |
| Location, *n* (%) | Proximal | 47 (26.9) |  |  |  |  |
|  | Distal | 128 (73.1) |  |  |  |  |

### Methylome profiling of colorectal tumors and identification of fecal DNA methylation markers

In order to identify effective methylation markers for CRC, we performed comprehensive methylome profiling of CRC by applying whole genome bisulfite sequencing (WGBS) to paired colorectal cancer tissues and matched normal tissue adjacent to tumor (NAT) samples (n=38). As expected, we found that CRC tumor tissues displayed distinct methylation patterns from NAT tissues (Fig.1A). By applying Least Absolute Shrinkage and Selection Operator (LASSO) algorithm, a total of 16277 DMLs in 1453 genes were identified, most of which were hyper-methylated (increased methylation in cancer tissue *vs*. normal; hyper-DMLs) (Fig. 1B). The top 64 DMLs distributed in 19 genes were selected as candidate targets for quantitative methylation-specific PCR (qMSP) and assays were sequentially validated using fully methylated and un-methylated standards, CRC and NAT tissue samples, and a small number of fecal DNA samples from patients and controls (Fig. 1C). Primer and probe pairs with satisfactory amplification profiles were then selected for further analysis.

**Fig 1.**
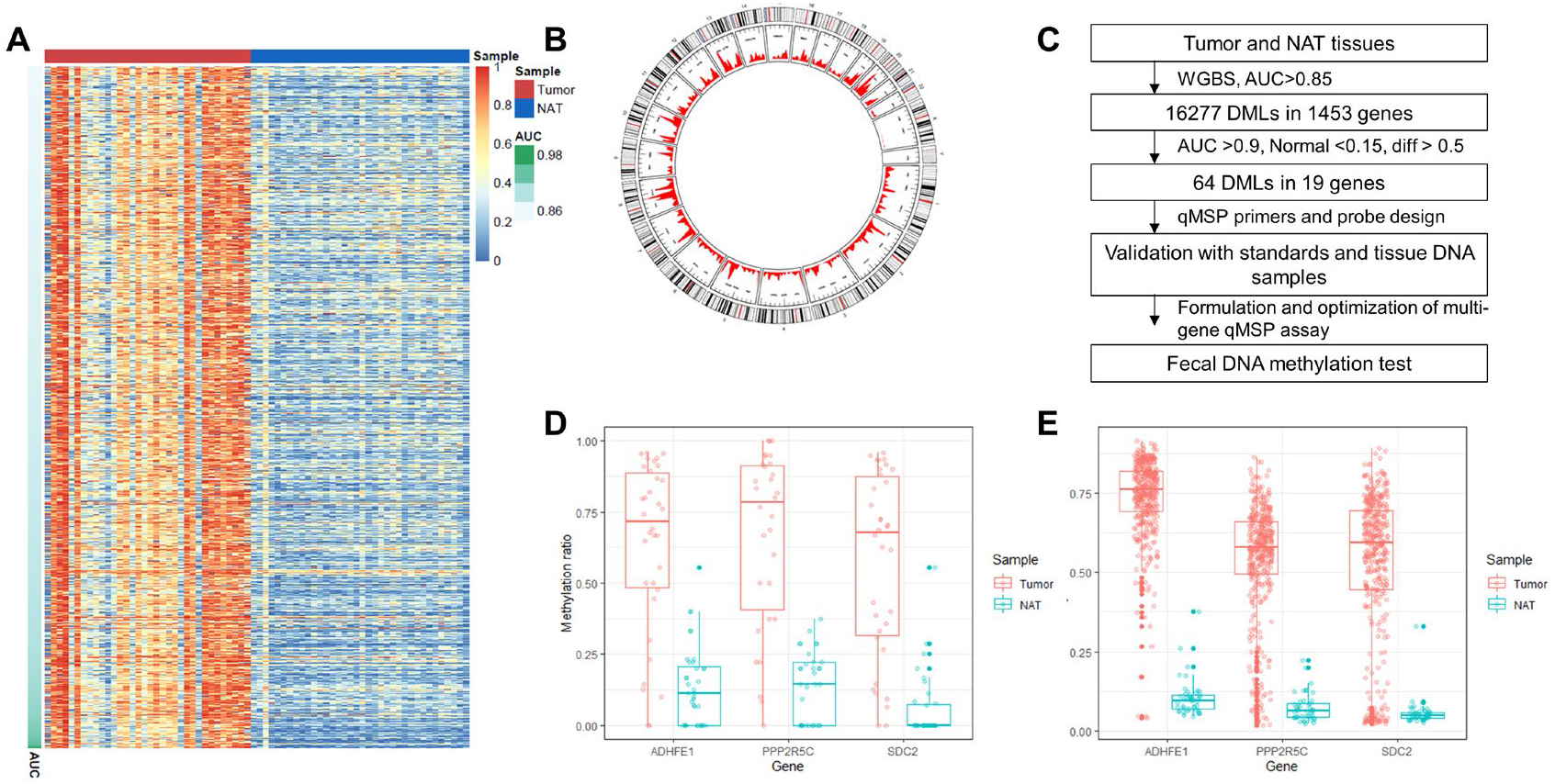
Identification of CRC-specific DMLs and Fecal DNA Methylation Markers for Non-invasive Detection of CRC. **A**. Heatmap of methylation levels of DMLs in CRC tumor and NAT tissues, ordered by AUC of each individual DML-based classfication model for differentiating CRC from NAT tissue. Block color represents the methylation level of each DML. **B**. Circos plot depicting the distribution of DMLs in human genome. Red points: Hyper-methylated DMLs (CRC tissues *vs*. NAT tissues). Blue points: hypo-methylated DMLs. From outer to inner circle are: overview of DMLs, area statistics of hyper-methylated DMLs, area statistics of hypo-methylation DMLs. **C**. Diagram depicting the process of marker selection and validation, as well as development of multiplex qMSP assay. **D and E**. The methylation levels of *SDC2, ADHFE1* and *PPP2R5C* in tumor and NAT tissues according to WGBS data (**D**) as well as The Cancer Genome Atlas (TCGA) database (**E**). For CpG sites that were not included in the TCGA database, methylation levels of nearby loci were shown.

Using this approach, three methylation markers (*SDC2, ADHFE1* and *PPP2R5C*) were found to show consistently higher methylation signal in CRC tissues *vs*. NAT and the same trend was also confirmed using the TCGA 450K BeadChip data (Fig. 1D,E); preliminary validation of each of the three markers using a subset of the fecal DNA samples also demonstrated expected amplification patter (amplification signals in most CRC stools and little to no amplification in controls; data not shown). The syndecan 2 gene (*SDC2*) encodes an intrinsic membrane protein that is involved in cell division, cell migration, and interstitial interactions^36, 37^. Various studies have shown that methylation level of *SDC2* was consistently higher in colorectal cancer tissues than adjacent normal tissues and could serve as an effective marker for the stool DNA based non-invasive diagnosis of colorectal cancer^19^. *SDC2* was re-discovered in our cohort as showing significantly differential methylation level between CRC and NAT tissues and demonstrated clear diagnostic power as stool DNA-based biomarker. The alcohol dehydrogenase iron containing 1 gene (*ADHFE1*) encodes a transhydrogenase that is involved in intracellular metabolism and other physiological activities. *ADHFE1* has been previously reported to be hypermethylated in advanced adenoma and carcinoma tissues ^38, 39^, and hypermethylation of *ADHFE1* had been shown to promote the proliferation of colorectal cancer cell via modulating cell cycle progression^40-42^. The Protein Phosphatase 2 Regulatory Subunit BGamma (*PPP2R5C*) gene encodes a regulatory subunit of the PP2A phosphatase, which dephosphorylates and activates TP53 and plays a role in response to DNA damage^43^. *PPP2R5C* has been shown to function as a tumor suppressor and inhibits colorectal cancer cell proliferation^44^. It has been suggested that hypermethylation of *PPP2R5C* gene is closely related to colorectal tumorigenesis as well as progression and may serve as a marker for detecting colorectal cancer^45^.

### Development and performance validation of a multi-gene fecal DNA methylation test

Considering tumor heterogeneity, we hypothesized that by combined detection of two or more of these methylated marker genes, we may be able to increase the chance to detect malignant and pre-malignant lesions; therefore, multiplex assays were established by combining these three genes in one qMSP assay, using *GAPDH* as the internal control gene (see Methods for details).

We went on to evaluate the diagnostic performance of the multiplex qMSP assay with fecal DNA samples. A total of 486 stool samples including CRC, APL, and control subjects (including NAN subjects, and subjects with negative findings on colonoscopy) were analyzed. We confirmed that methylation levels of all 3 markers measured by ΔCt (Ct value of methylated target gene minus that of internal control gene) were significantly higher in stool samples from CRC and APL than in non-advanced neoplasm subjects and subjects with negative findings on colonoscopy (Fig. 2A; p < 0.001). Samples were then divided into training and testing set with a 6:4 split, and random forest models were fitted using ΔCt values of single or combined markers as features. The result showed that better performance (an AUC of 0.943 and 0.927 in the training and testing set, respectively) was achieved when all three markers were used as features (Figure 2B, C). In order to develop a clinically applicable algorithm to classify CRC *vs*. control, we subsequently selected the optimal threshold for each marker gene based on Youden index and applied a simple OR logic (sample is predicted positive when either marker gene tests positive) to integrate the results of these three markers (supplementary Table 1). We found that such assay provided robust classification performance: in the training set, it generated a sensitivity of 88.6% for CRC cases and a specificity of 93.2% for control subjects; in the testing set, a sensitivity of 87.1% for CRC subjects and a specificity of 89.8% for the controls were achieved. In the testing set, the assay detected 46.2% (12/26) of APLs, and 18.2% (4/22) of other cancers were tested positive (Fig 2D). We also investigated the diagnostic efficacy of the multi-gene methylation assay among CRC patients stratified by AJCC stages and tumor locations. It showed a sensitivity of 93.0% and 74.5% for distal CRC (including descending, sigmoid colon, and rectum) and proximal CRC (from the appendix to splenic flexure), respectively. It also detected more curable AJCC stage (stage I-III) CRCs than stage IV CRCs (86.5%, 88.4%, 92.6% vs. 73.3%).

**Fig 2.**
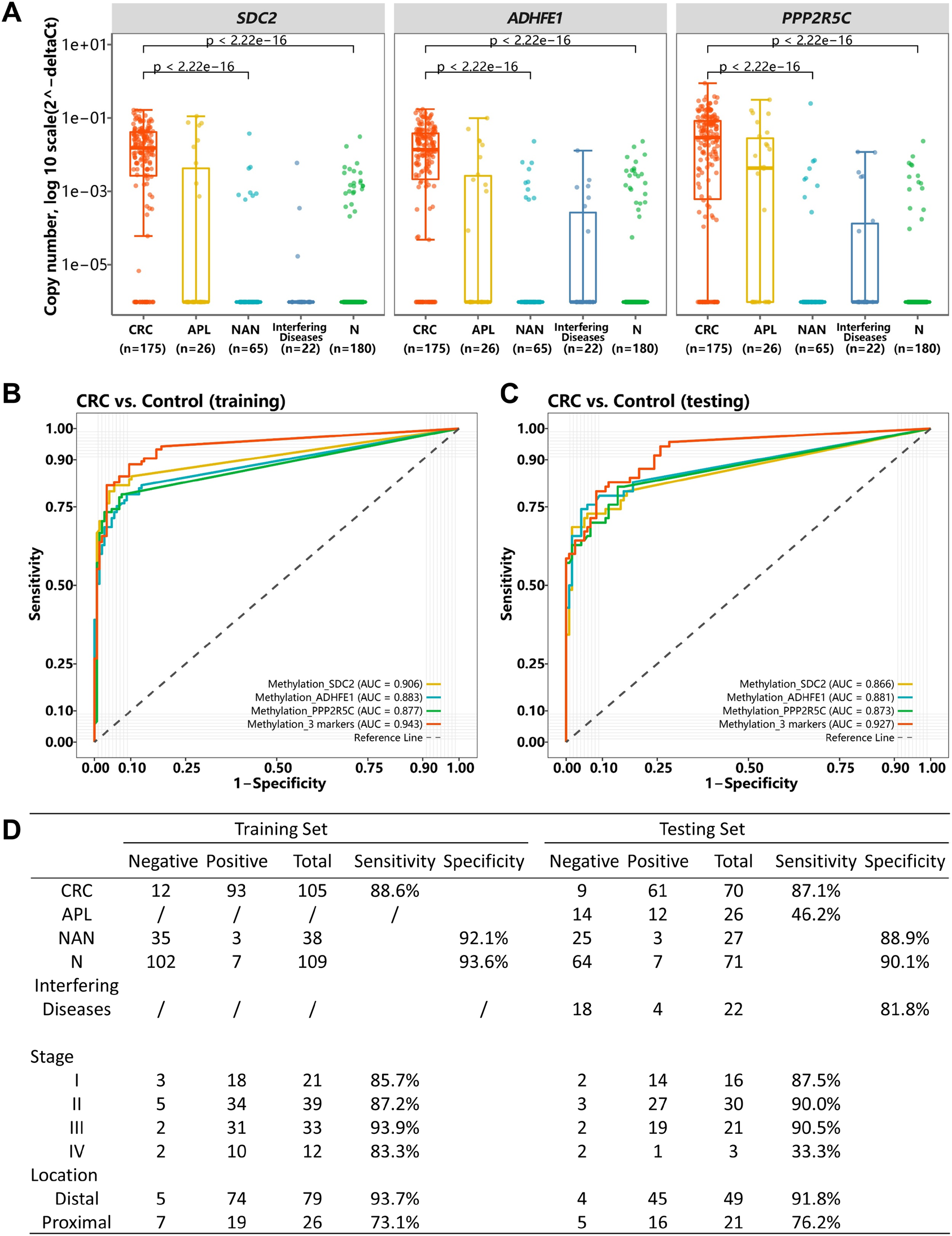
The performance of three methylation markers in detecting CRC in fecal DNA. **A**. Methylation copy numbers of selected methylation markers in fecal DNA samples. **B, C. Performance** of the methylation marker-based models for detecting CRC *vs*. controls in the training (B) and testing set (C), using either single marker or combination of all three markers. Controls included non-advanced neoplasm subjects and subjects with negative findings on colonoscopy. **D**. Diagnostic performance of multi-gene fecal DNA methylation test in training and testing set, stratified by AJCC stages and tumor locations.

### Fecal DNA mutation-based CRC diagnosis

To test whether genomic alterations of fecal DNA could be potentially used for CRC detection, we analyzed the fecal DNA mutational profile for subjects with enough stool sample available for additional analysis. A total of 155 CRC patients and 175 non-CRC subjects including APL (n=23), NAN (n=44) and subjects with negative colonoscopy results (n=108) were analyzed by targeted sequencing with a panel that targets exons of 139 cancer driver genes, predicted to have nearly 100% coverage for CRC cases according to TCGA data (supplementary Figure 1)^34^. In total, 233 mutations were detected in 64 (41.3%) CRC samples, two mutations were detected in two (1.3%) control samples, and one mutation was detected from one APL sample. *TP53* (22%), *APC* (21%) and *KRAS* (19%) were the most frequently mutated genes in fecal DNA (Fig. 3A). The overall mutation spectrum was highly consistent with TCGA data, showing high prevalence for variants in genes such as *APC, TP53, KRAS, PIK3CA, FBXW7*, and *BRAF*, etc (supplementary Figure 2). In CRC samples, distribution of mutation allele frequencies (AFs) showed no apparent trend from stage I to stage IV (Fig. 3B).

**Figure 3.**
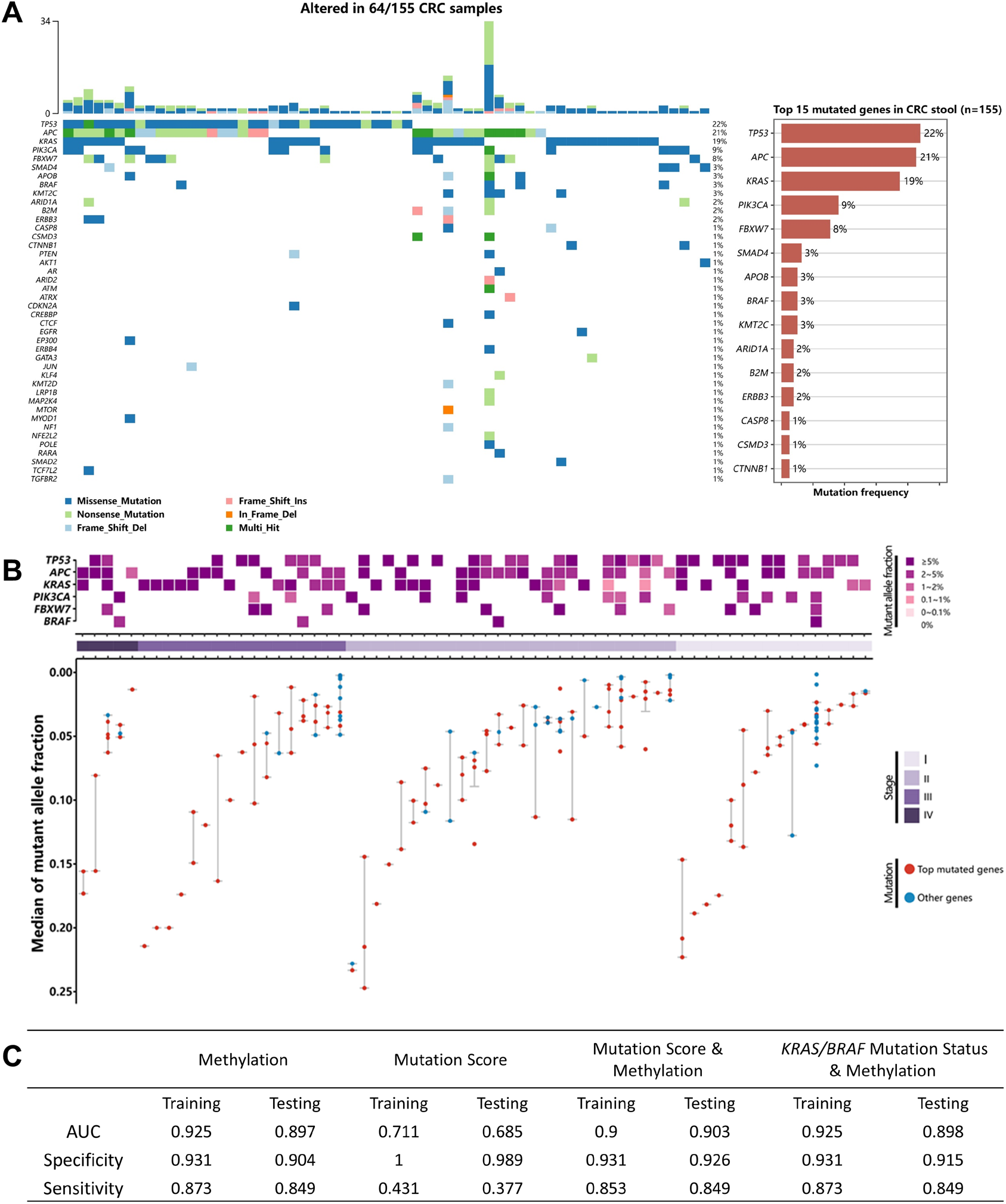
Identification and analysis of genomic alteration in fecal DNA. **A**. Mutational landscape of fecal DNA from CRC patients. Each column represents a CRC case. Upper bar chart represents the number of mutations in each sample. Lower waterfall diagram shows the mutations in each sample. Right bar plot represents top 15 mutated genes in CRC fecal DNA. **B**. Hotspot mutations identified from the CRC fecal DNA. **C**. Performance of the diagnostic models in the training and testing set using methylation-based model, mutation-based diagnostic model or integrated model that combines both mutation and methylation status.

We fitted a mutation-based diagnostic model based by calculating a weighted mutational burden (mutation score) using previously described approach^34^ (see methods for details). The model showed superior specificities compared to the methylation markers-based model (100% in training set and 98.9% in testing set); however, the sensitivities was much lower for detecting CRCs (43.1% in training set and 37.7% in testing set) and APLs (4.3% in the testing set). Moreover, we found that the diagnostic performance was not improved by combining the methylation model with the mutation-based model, when either mutation scores or status (presence/absence) of hotspot mutations in gene *KRAS/BRAF* were used as features.

### Stool microbiome-based CRC diagnosis

To test whether stool microbiome could be used as a tool for CRC detection, microbiome signatures of CRC, APL, and control fecal DNA were analyzed by metagenomics sequencing (Figure 4A). Among the 468 fecal samples included in methylation qPCR analysis, a total of 327 samples (155CRC, 23APL and 149 non-CRC/APL control samples were analyzed. We developed a random forest classification model for CRC *vs*. Controls based on the relative abundance of microbial species, which showed an AUC of 0.91 in training set and 0.83 in testing set, a performance slightly inferior to the multi-gene methylation assay (Figure 4B,C). With this model, advanced precancerous lesion samples were more likely to be predicted as positive than non-CRC/APL controls (supplemental Figure 3), suggesting that advanced precancerous lesions start to enrich for CRC-associated microbial signatures during the transition from the benign to the malignant state.

**Fig.4.**
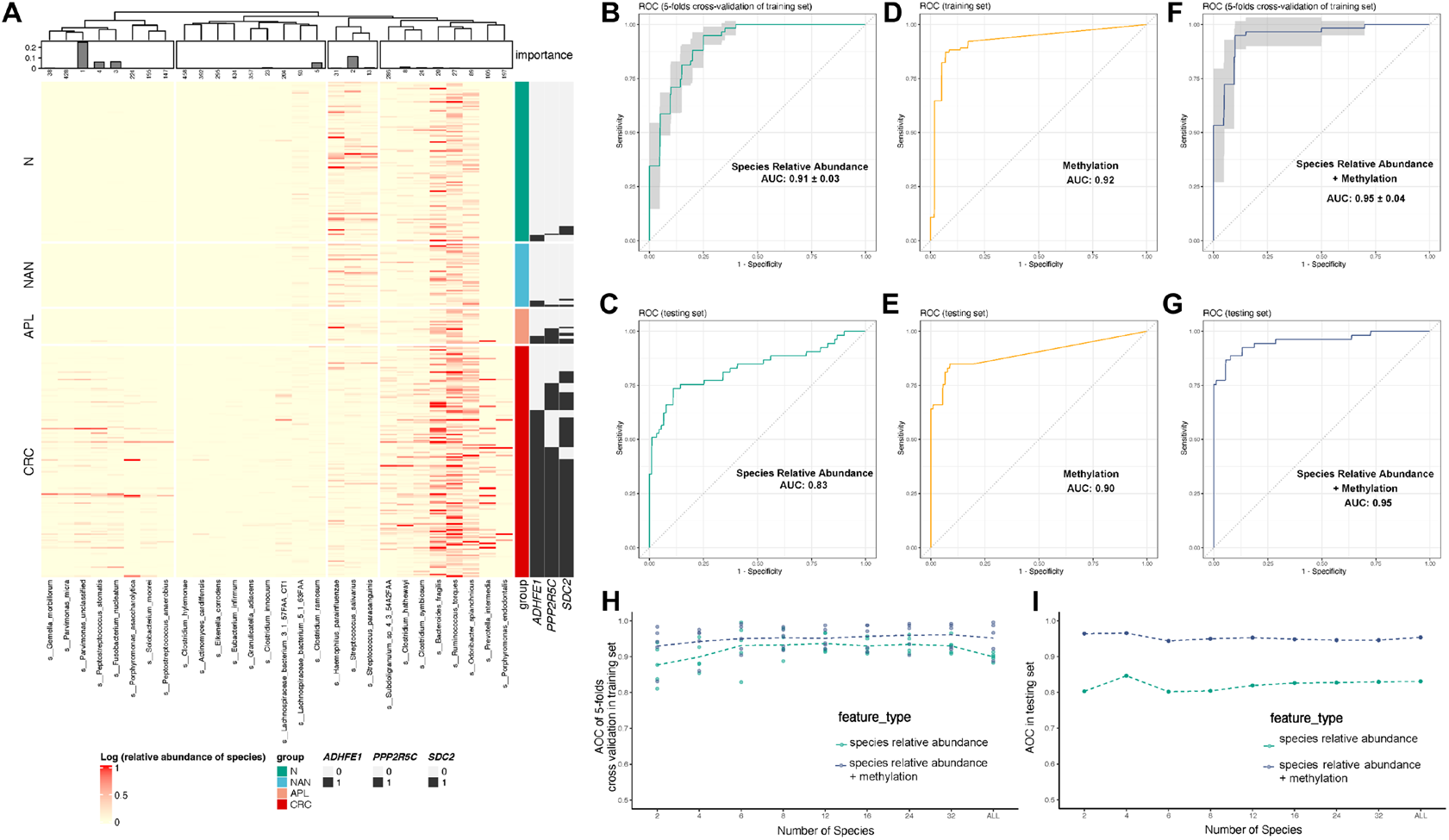
Performance of gut microbiota-based and integrated multi-target diagnostic models. **A**. Heatmap shows relative abundance of marker microbial species in stool samples of CRC and control. The marker species were detected by linear decomposition model (LDM), adjusted by age, gender, filtered by FDR = 0.01. Samples were ordered by the multi-gene methylation test result, and species were clustered based on the Bray-Curtis distance of all samples. Bar plot on the top: Y-axis represents feature importance in the random forest model; X-axis represents rank of features. **B-G**. ROC curves of microbiota-based model (**B, C**), methylation-based model (**D, E**), and integrated model (**F, G**). Controls included non-advanced neoplasm subjects and subjects with negative findings on colonoscopy. **H, I**. Performance of the microbiota-based model and the integrated model with deceasing number of features.

Because the cost of WGS-based metagenomics assay is relatively high due to sequencing cost, it would be worthwhile to identify a handful of microbial clade markers in order to develop a qPCR-based assay. Several microbial species showed outstanding feature importance in our random forest model, such as those that appeared to be consistently enriched in CRCs *vs*. controls including *Fusobacterium nucleatum, Parvimonas unclassified*, and *Parvimonas micra*, as well as *streptococcus salivarius*, which appeared to be enriched in non-CRC stool samples (Figure 4A). To test the feasibility of utilizing a small number of marker species for development of a diagnostic assay, we performed feature selection by feature elimination which showed that the CV AUC largely remained stable when we reduced the full set of features down to only two species (varied within the range of 0.88 to 0.91; Figure 4H). In the testing set, the model also showed a relative stable AUC varying between 0.8 to 0.85 when feature number was reduced to two (Figure 4I).

Previous studies have applied metagenomics sequencing to stool samples of CRCs and controls to discover CRC-associated microbial signatures^46-50^. To test whether the pattern discovered from our metagenomics data is consistent with previous findings and/or our diagnostic model could be generalized to independent cohorts, we tested our model on previous published microbiome datasets as independent testing set and found that it achieved a pooled AUC of 0.76 (supplementary Figure 4A), slightly inferior than the performance in our testing set. Also, when decreasing the feature number to just a couple, the models demonstrated lower accuracy in public datasets (supplementary Figure 4B), a trend recapitulating the observation by Thomas et al., suggesting that it would be challenging to develop a multiplex qPCR-based diagnostic assay based on just a few microbial species markers. Collectively, these results demonstrated the feasibility of gut microbiome in detecting colorectal cancer and precancerous lesions. However, the overall diagnostic accuracy appeared to be inferior to what could be achieved by just a few methylation markers.

### Integrated diagnostic model combining methylation and metagenomic markers

Since our results suggest that microbial markers-based classification model had slightly inferior accuracy compared to methylation markers-based assay, we went on to test whether integrated analysis of methylation markers and microbial markers may further enhance the diagnostic performance. To do this, we trained a random forest model on the multi-gene methylation data as well as relative abundance data of microbial species, which showed an improved CV AUC in the training set (0.95) as well as in the testing set (0.95) (Figure 4F, G). Remarkably, when number of features was reduced to just two species, the integrated model still showed a high accuracy in the testing set (AUC=0.96; Figure 4I). Taken together, our findings suggest that gut microbiome features may provide further improvement in model accuracy when combined with methylation-based markers and highlighted the potential of integrated multi-target fecal DNA assay for enhancing diagnostic performance for CRC.

## Discussion

In current study, we identified fecal DNA methylation markers for CRC detection by WGBS of CRC and normal tissues, and subsequently designed and validated qMSP assays of candidate markers in fecal DNA, focusing on their ability to detect CRC/APL *vs*. non-advanced neoplasm and normal colorectal epithelium. The final stool-based assay including a panel of three methylated gene markers *SDC2, ADHFE1*, and *PPP2R5C* achieved a sensitivity of 87.1% in detecting CRC, 46.2% for APL, and had a specificity of 89.8% for controls including negative results on colonoscopy, non-advanced neoplasms and other lesions in testing set. We noticed that the assay had a higher sensitivity in detecting distal CRC than proximal CRC, an observation consistent with previous reports^18, 36^, and one explanation might be that the more hydrated environment in the proximal colon is more hostile towards DNA preservation^51^.

Previously, it was reported that integration of methylation markers, *KRAS* mutation, and FIT overall provided optimal diagnostic performance for detection of CRC and APL versus control populations, but the relative contribution of each type of biomarker remains unclear. Here, we found that genomic alterations overall provided inferior performance compared to the panel of methylation markers, and importantly, our attempt to integrate the mutation profile with the methylation markers showed marginal enhancement. The mutational landscape of human genome identified in stool DNA was highly consistent with what has been known for Chinese CRC patients^52^, which suggests that these mutant genomes indeed came from malignant colorectal epithelial cells and mutational profile of fecal DNA may be used effectively as non-invasive markers for CRC. Nevertheless, our results showed that mutational status as diagnostic markers provided high specificity (near 100%) but sensitivity was suboptimal. In general, the most frequently mutated genes of CRC were also found to bear the most mutations in fecal DNA, albeit with lower detection rates compared to what would be expected for CRC tissue. For example, it has been shown that prevalence of somatic APC mutations is lower in Chinese patients compared to TCGA data (43.5% *vs*. 78.6%); here in current study, *APC* mutations were identified in ∼20% of CRC stool samples. Failure to detect all *APC* mutations from fecal DNA samples may be attributed to the limited amount of human DNA that can be recovered from stool as well as heterogeneity of stool samples, which may make detection of clonal mutations more difficult^53^. Also, mutations were almost never detected in stool DNA for APL cases. Although *APC*/*KRAS* mutations are believed to arise relatively early during CRC tumorigenesis (i.e., before or during the adenoma stage)^54^, the fact that we failed to detect their presence may be attributed to limited DNA shedding of APL, which also generally have relatively smaller tumor mass.

Meanwhile, by comparing the metagenomics data of CRC versus control cases, we were able to identify gut microbiota species that showed differential abundance between the two groups. Diagnostic model based on relative abundance of bacterial species exhibited overall inferior diagnostic performance in differentiating CRC from non-CRC/APL controls compared to methylation-based test. Out of the species that showed top feature importance in the microbiome-based classification models, most showed notably higher abundance in CRC stool samples *vs*. control, such as *Fusobacterium nucleatum, Peptostreptococcus stomatis, and Parvimonas unclassified*. Importantly, these species have all been reported to be markedly upregulated in CRC stools^47, 55^. *streptococcus salivarius*, on the other hand, is a probiotic associated with inhibition of chronic inflammation as well as tumorigenesis^56^. By meta-analysis, Thomas et al. identified cross-cohort microbial diagnostic signatures, with the most significant CRC-associated features being *F. nucleatum, Parvinomas micra, Gemella morbillorum, P. stomatis, and Solobacterium moorei*, all of which also showed significant enrichment in CRC group in our cohort. It was suggested that microbial signatures can be highly variable and overall could show poor transportability from study to study due to difference in experimental conditions and/or population characteristics^57^. Indeed, we found that using public datasets as independent validation set, the microbiota-based model mostly showed slightly inferior performance than in our testing set, with an AUC ranging from 0.71 to 0.86 (Supplementary Figure 4A). Nevertheless, the identified CRC-enriched species from our cohort were still highly consistent with previous findings, highlighting the potential diagnostic values of these microbial markers, which may serve as a universal tool for CRC detection across different ethnic/genetic background and geological regions.

Previous study showed that combination of multiple analytes in stool might further enhance diagnostic performance for CRC^22^. Therefore, after showing that mutation or metagenomics markers both provided inferior performance than methylation markers, we went on to test whether combination of these markers may further improve the diagnostic accuracy. Our results showed that targeted mutation profiling or detection of hotspot mutations appeared to add little to the diagnostic performance of the methylation markers. Yet, metagenomic markers appeared to have the potential to further enhance the diagnostic performance, even when utilizing just a few (i.e., two to three) of microbial species as features along with the panel of methylation markers. Taken together, these findings highlight the prominent diagnostic power of methylation markers and suggest that the methylation test alone may serve as a promising tool for non-invasive CRC diagnosis and possibly CRC screening, which may be further enhanced by a multi-target fecal DNA assay that combines the detection of methylated human DNA and microbial genomes in the stool.

We notice a couple of limitations in current study. First, study subjects were from a single study site, which may potentially have caused bias in reported data. A larger scale, multi-center study would be needed to fully evaluate the clinical performance of the stool DNA methylation-based test and the combined detection of methylation and microbial markers. Secondly, we only included a limited number of advanced pre-cancer malignancies such as APL patients in the current study, and we mostly focused on identifying and validating markers for effective detection of CRC. In the future, it may also be worthwhile to specifically look for markers that could provide enhanced sensitivity for advanced pre-cancer malignancies.

Despite these limitations, the present study was the first one to our knowledge that comprehensively investigated fecal DNA methylation, mutation profile, and gut microbiome in the same cohort and compared their values for CRC diagnosis. Overall, our results indicated that methylation markers can provide superior diagnostic performance and highlighted that multi-target detection approach such as integration of methylation and microbial markers may further improve the performance. Further efforts would be needed to develop and optimize such an assay. Our findings provided foundation for future investigations towards optimization of fecal DNA based diagnostic and screening tools that may help improve prevention and early intervention of CRC.

## Data Availability

All data produced in the present study are available upon reasonable request to the authors

## Acknowledgements

We are indebted to Tao Zhang and Kaiye Cai for valuable discussions regarding the microbiota-based diagnostic model.

