## Supplementary Table & Figures for "Identification of multi-omic biomarkers from Fecal DNA for improved Detection of Colorectal Cancer and precancerous lesions"

Supplementary Table 1. Diagnostic Threshold of the Multi-gene Methylation Test.

Supplementary Figure 1. Percentage of TCGA samples predicted to be covered by the targeted sequencing panel utilized in the present study by different cancer types.

Supplementary Figure 2. Comparison of mutational landscape of fecal DNA with TCGA tissue data for CRC patients.

Supplementary Figure 3. Predicted probabilities of CRC status by fecal microbiota-based model and integrated model that combines fecal methylation status and microbiota data in different study groups.

Supplementary Figure 4. Performance of fecal DNA microbiota-based model in previously published stool microbiome datasets.

**Supplementary Table 1. Diagnostic Threshold of the Multi-gene Methylation Test.**

| Ct value of GAPDH | Target gene | The Ct value of target gene ① | The △Ct of target gene ② | Result |
|---|---|---|---|---|
| Ct≤37 | SDC2 | Ct≤38 | ≤9 | A sample tests "POSITIVE" if the result of one or more genes satisfies both condition ① and ②.<br>In any other cases, a test sample is "NEGATIVE". |
| | ADHFE1 | Ct≤38 | ≤8 | |
| | PPP2R5C | Ct≤38 | ≤7 | |
| Ct>37 | / | Any value | Any value | INVALID, retest the sample |

*△Ct: △Ct = (the Ct value of target gene) – (the Ct value of GAPDH).

**Supplementary Figure 1. Percentage of TCGA samples predicted to be covered by the targeted sequencing panel utilized in the present study by different cancer types.**

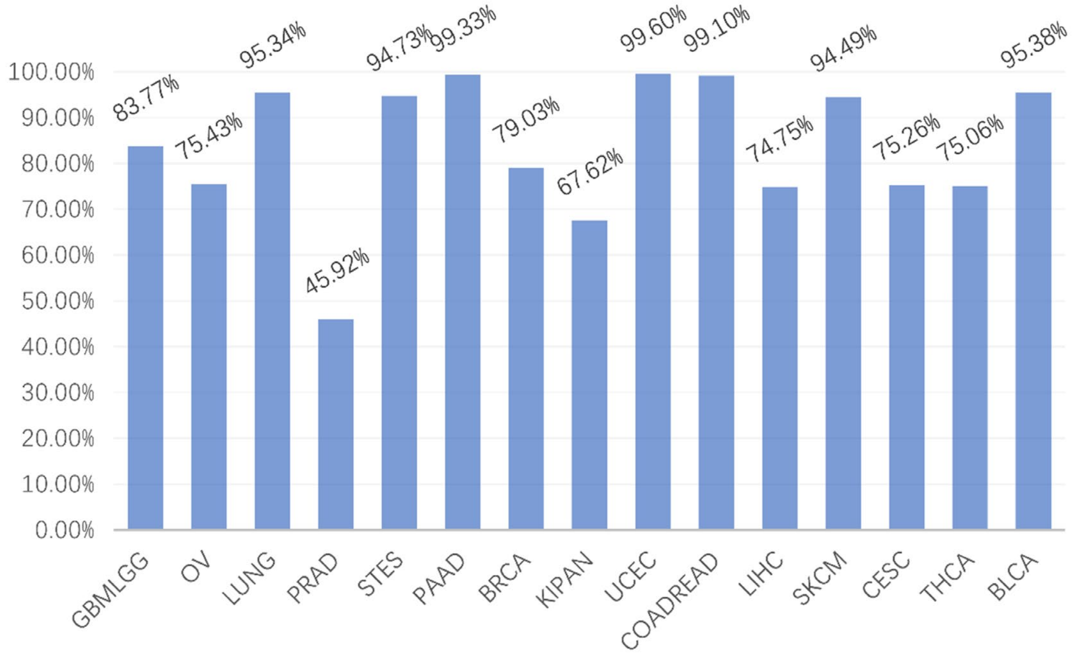


GBMLGG: Glioblastoma multiforme and Brain Lower Grade Glioma; OV: Ovarian serous cystadenocarcinoma; LUNG: Lung adenocarcinoma; PRAD: Prostate adenocarcinoma; STADES: Stomach adenocarcinoma and Esophageal carcinoma; PAAD: Pancreatic adenocarcinoma; BRCA: Breast invasive carcinoma; KIPAN: Kidney pan- cancer; UCEC: Uterine Corpus Endometrial Carcinoma; COADREAD: Colon adenocarcinoma and Rectum adenocarcinoma; LIHC: Liver hepatocellular carcinoma; CESC: Cervical squamous cell carcinoma and endocervical adenocarcinoma; THCA: Thyroid carcinoma; BLCA: Bladder Urothelial Carcinoma

**Supplementary Figure 2. Comparison of mutational landscape of fecal DNA with TCGA tissue data for CRC patients.**

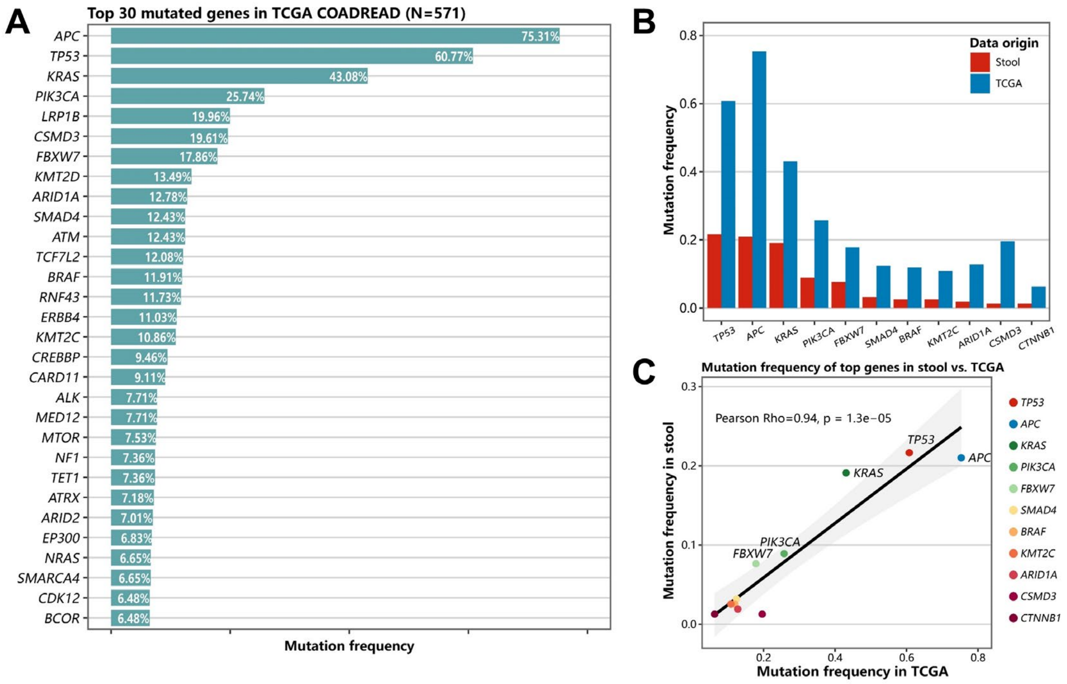


**A.** Mutation frequency of top mutated genes in TCGA CRC tissue data. **B.** Comparison of mutation frequencies of top mutated genes identified in our study with TCGA data. **C,** Correlation of mutation frequency of top mutated genes with TCGA data.

**Supplementary Figure 3. Predicted probabilities of CRC status by fecal microbiota-based model and integrated model that combines fecal methylation status and microbiota data in different study groups.**

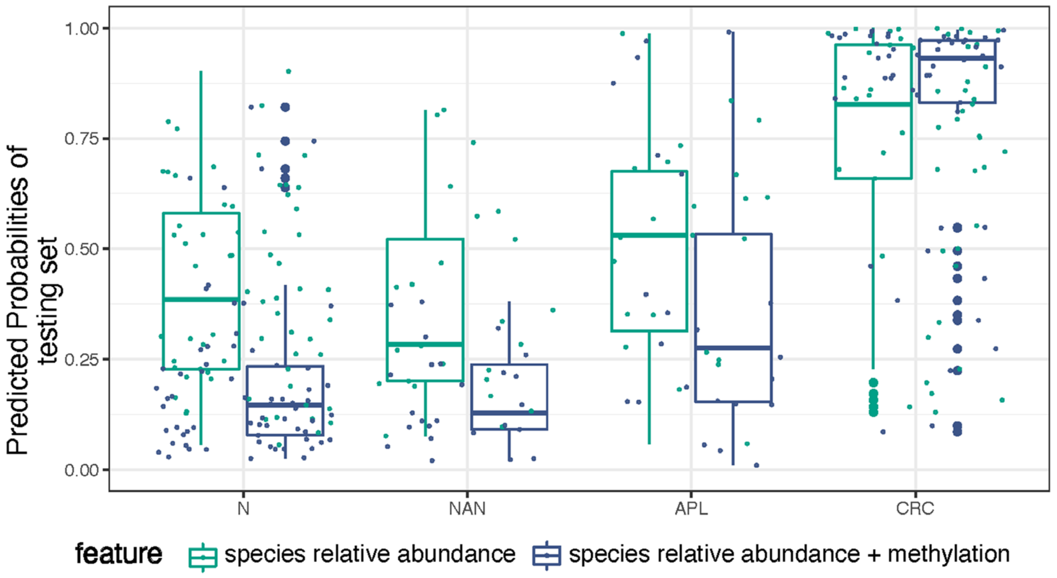


N, normal; NAN non-advanced neoplasm; APL, advanced precancerous lesions; CRC, colorectal cancer.

**Supplementary Figure 4. Performance of fecal DNA microbiota-based model in previously published stool microbiome datasets.**

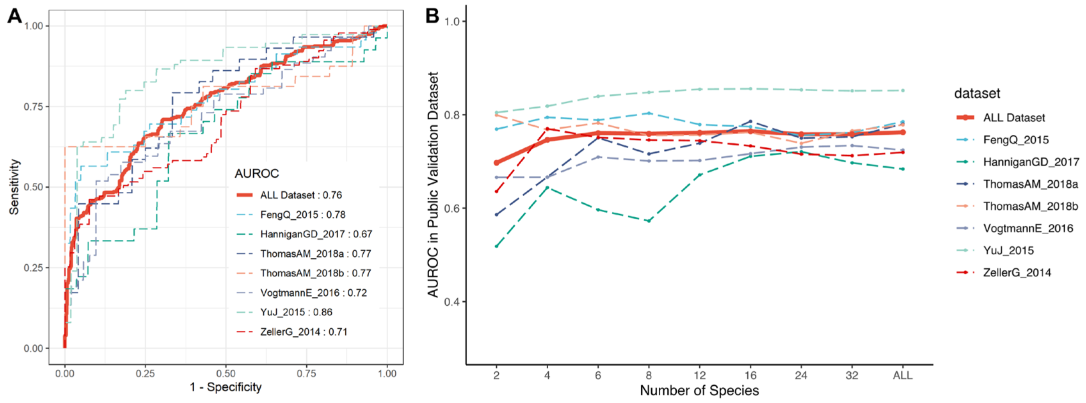


**A.** ROC curves of fecal microbiota-based model in previously published stool microbiome datasets.
**B.** Model performance with deceasing number of features published datasets.
